# Prenatal Exposure of Pesticide Mixtures and the Placental Transcriptome: Insights from Trimester-specific, Sex-Specific and Metabolite-Scaled Analyses in the SAWASDEE Cohort

**DOI:** 10.1101/2024.09.16.24313768

**Authors:** Yewei Wang, Karen Hermetz, Amber Burt, Corina Lesseur, Parinya Panuwet, Nancy Fiedler, Tippawan Prapamontol, Panrapee Suttiwan, Pimjuta Nimmapirat, Supattra Sittiwang, Warangkana Naksen, Volha Yakimavets, Dana Boyd Barr, Ke Hao, Jia Chen, Carmen J. Marsit

## Abstract

We investigated the effect of exposure to pesticide mixtures during pregnancy on the placental transcriptome, to link these exposures and placental functions. The Study of Asian Women and their Offspring’s Development and Environmental Exposures (SAWASDEE) enrolled pregnant farmworkers from Thailand (n=248), who were primarily exposed to organophosphate (OP) and pyrethroid pesticides. We measured maternal urinary levels of six non-specific OP metabolites expressed as three summary measures (dimethylalkylphosphates (DMAP), diethylalkylphosphates (DEAP), and dialkylphosphates (DAP) and three pyrethroid metabolites (3-phenoxybenzoic acid (3-PBA), *cis*- and *trans*-3-(2,2-dichlorovinyl)-2,2-dimethylcyclopropane carboxylic acid (Cis-DCCA, Trans-DCCA) during early, middle, and late pregnancy, and adjusted for urine dilution using creatinine. RNA-sequencing was used to profile the placental transcriptome from which 21 co-expression network modules were identified by Weighted Gene Co-expression Network Analysis. Quantile g-computation analysis identified an average pregnancy positive mixture exposure effect on the E2f Target Module (β = 0.013, p = 0.012) and a negative mixture exposure effect (β = −0.016, p = 0.008) on the Myogenesis Module. The pesticide metabolites driving the associations differed for each module, highlighting differential susceptibilities within the placental transcriptome to various pesticides. When stratifying by infant sex, the average pregnancy mixture exhibited a significant negative effect (β= −0.018, P=0.016) on the Myogenesis Module only in females; other modules, such as epithelial-mesenchymal transition, though not demonstrating an overall mixture effect, did demonstrate differential impacts of the mixture by sex. Pesticide mixtures in both the second trimester (β = −0.013, p = 0.015) and the third trimester (β = −0.012, p = 0.028) exhibited consistently significant negative associations with the Myogenesis module. These findings underscore the importance of considering the prenatal environment more holistically, understanding the placenta’s susceptibility to contaminants, and incorporating sex-specific analyses to understand differential impacts.

## Introduction

Pesticides, including the organophosphate and pyrethroid classes, are an integral part of modern agriculture worldwide, playing a crucial role in pest control and crop yield sustainability ^1,2^. Human exposure to these chemicals occurs through diverse pathways, including agricultural activities, residential applications, and dietary intake ^3^. As insecticides, organophosphates bind to acetylcholinesterase, causing an accumulation of acetylcholine at nerve synapses, while pyrethroids interfere with sodium channel functions, crucial for nerve signal transmission ^4^. The pervasive use of these pesticides, however, has raised significant concerns regarding their extensive human exposure ^5^. Despite their efficacy in targeting insect populations, mounting evidence suggests that these chemicals pose considerable neurotoxic risks to wildlife and humans, highlighting broader ecological and health implications ^6,7,8^.

The vulnerability of certain populations, especially pregnant women, is of particular concern ^9–11^. Both organophosphate and pyrethroid pesticides are lipophilic, enabling them to cross the placenta and the blood-brain barrier, thereby potentially disrupting critical periods of placenta and brain development ^12–15^. Given that many pesticides can traverse these biological barriers, prenatal exposure is worrisome, as the entire developmental system is exceptionally susceptible during gestation and early life. The placenta, essential for nutrient and gas exchange between mother and fetus, also serves as a mediator for fetal exposure to external toxins, including pesticides. Understanding how pesticide exposure influences placental function is critical, given its direct impact on developmental trajectories and long-term health outcomes in children ^16–20^.

The growing body of research on pesticide exposure during pregnancy highlights significant risks particularly through alterations in placental function ^21,22^. Recent studies have provided insights into how single classes of pesticides, such as organophosphates and pyrethroids, can alter placental signaling pathways. Studies have historically shown how single pesticides influence biomarkers like acetylcholinesterase and catalase activities or cause morphological changes in the placenta ^16,23–25^. Recent findings from our group have demonstrated that urinary metabolite levels of specific organophosphate or pyrethroid pesticides are associated with variation in placental gene modules representing functional changes to the placental transcriptome ^21,26,27^. For instance, urinary levels of the organophosphate pesticide metabolite diethylphosphate (DEP) have been linked to placental gene modules negatively associated with myogenesis and epithelial-to-mesenchymal transition (EMT) in late pregnancy while urinary levels of diethylthiophosphate (DETP) have been linked to hypoxia during early pregnancy ^26^. Similarly, urinary levels of pyrethroid metabolites, such as 3-phenoxybenzoic acid (3-PBA), cis-3-(2,2-Dichlorovinyl)-2,2-dimethylcyclopropane carboxylic acid (cis-DCCA), and trans-3-(2,2-Dichlorovinyl)-2,2-dimethylcyclopropane carboxylic acid (trans-DCCA), have been linked to changes in the placental transcriptome ^27^. Notably, these metabolites affect genes involved in oxidative phosphorylation and immune response pathways ^27^. These findings underline the placenta’s sensitivity to chemical exposures and thus potential health risks to fetuses and offspring.

While these studies demonstrate the effects of individual classes of pesticide metabolites, a significant knowledge gap remains regarding the consequences of simultaneous exposure to multiple classes of pesticides that affect neurons^28–31^. Such mixed exposures more accurately mirror the complex environmental conditions commonly encountered in agricultural settings and involve intricate interactions whose combined impacts on health are not yet fully understood. Research by Acosta-Maldonado and Tyagi suggests that exposure to mixed pesticides may accelerate placental maturity and are correlated with adverse birth outcomes, including preterm labor ^16,32^. The cumulative effects of these mixed exposures on placenta, especially in terms of how these compounds interact to affect gene expression within the placenta, are poorly documented ^33–36^. Bridging this gap is crucial for developing more effective public health strategies and protective measures for pregnant women and their developing fetuses, as understanding these interactions could lead to beter risk assessment and intervention methods in environments with high pesticide use.

By incorporating maternal prenatal urinary metabolites derived from exposure to multiple pesticides into a quantile g-computation mixture analysis, this study aims to provide a more comprehensive representation of the complex exposure scenarios commonly faced by pregnant women in agricultural communities from the SAWASDEE cohort. This approach allows us to dissect the complex associations of mixture prenatal pesticide exposures on placental gene expression across different stages of pregnancy and between male and female fetuses, enhancing our understanding of the interplay between environmental exposures and placental functions.

## Methods

### Study Design and Population

This study analyzed the relationship between prenatal urinary pesticide metabolite levels and placental transcriptomics, building on the foundation laid by the SAWASDEE study which focused on agricultural workers in Thailand from 2017 to 2019 ^37^. Pregnant women were recruited during their first trimester from community health clinics and district hospitals within Chiang Mai province, encompassing two agriculturally intensive districts. Non-singleton pregnancies or those with major complications potentially affecting fetal growth were excluded. Eligibility criteria are included in the published protocol ^37^. A cohort of 394 pregnant women was initially recruited. Of these, 333 completed the pregnancy with the delivery of live infants. Pesticide exposure assessments were successfully conducted in 332 of these women. From the cohort with live births, 253 participants provided placental samples that met our criteria for transcriptomic analysis ^26^. After accounting for overlap in data availability and quality, our final sample size consisted of 248 mother-infant pairs.

### Prenatal Pesticides Exposure Assessment

Multiple maternal urine samples were collected across three key stages of pregnancy—early (0–14 weeks gestation), middle (14 weeks 1 day–27 weeks gestation), and late (more than 27 weeks gestation)—as part of a composite sampling strategy to characterize exposure to both pyrethroid and organophosphate pesticides throughout pregnancy ^37^. The samples were collected in 100 mL polypropylene containers and aliquoted. Composites were created using equal volumes from each sample collected during the same pregnancy period, ensuring that these composites represented the average of the analyte concentrations for each period. For pyrethroids, urinary levels of 3-phenoxybenzoic acid (3-PBA) and cis- and trans-3-(2,2-dichlorovinyl)-2,2-dimethylcyclopropane carboxylic acid (cis-DCCA, trans-DCCA) were measured in the samples using liquid chromatography-tandem mass spectrometry (LC-MS/MS) with negative-mode electrospray ionization (ESI) and isotope dilution quantitation ^27^. The quality of the analysis was ensured by semi-annual German External Quality Assessment Scheme (GEQUAS) certification and the inclusion of NIST SRM 3667 standards. For organophosphates, six dialkylphosphate metabolites were measured in Thailand using gas chromatography-flame photometric detection (GC-FPD), validated against mass spectrometry-based methods. However, due to high rates of non-detection, only diethyl phosphate (DEP) and diethyl thiophosphate (DETP) remained in further analyses. For values below the limits of detection (LOD), concentrations were imputed using the LOD divided by the square root of 2 ^38^. Quality control included analysis of blank and spiked samples within each analytical run to ensure the integrity of the data, supported by successful participation in the GEQUAS. In both analyses, the concentrations of the metabolites were normalized using urinary creatinine measurements, which were conducted by spiking the urine samples with isotopically labeled analogues and diluting them 1000-fold prior to analysis using LC-MS/MS ^39^. This normalization accounts for variations in urine dilution across different samples. The distributional data for these metabolites have been previously reported in earlier publications ^26^ ^27^.

### Placenta collection and RNA extraction

The procedure for placental collection and subsequent RNA extraction adhered to the methodologies outlined by Baumert ^37^. Within a post-delivery window of two hours, five circular tissue slices, each approximately 1 inch in diameter, were excised from placental areas devoid of maternal decidua and situated approximately 2 cm from the umbilical cord insertion site. To prevent RNA degradation, these samples were immediately rinsed to remove any contaminating blood or debris and submerged in RNALater solution (Invitrogen), an RNA stabilization agent. After ensuring stabilization in RNALater for no less than 72 hours at a controlled temperature of 4°C, the samples were rapidly frozen using liquid nitrogen then homogenized. These homogenized samples were then transferred to 2 mL cryovials and securely stored at −80°C to maintain RNA integrity until analysis was initiated. RNA extraction was conducted using the Norgen Animal Tissue RNA Purification Kit, selected for its efficacy in extracting high-quality RNA from tissue matrices. The concentration and purity of the extracted RNA were quantitatively assessed using a NanoDrop 2000 spectrophotometer, and the RNA’s structural integrity was further verified via gel electrophoresis using an Agilent 2100 Bioanalyzer.

### Placental Transcriptome Profiling

Transcriptome profiling followed the method described in the previous publication ^26^. Initially, RNA sequencing (RNA-Seq) libraries were prepared, starting with the depletion of ribosomal RNA (rRNA) using the RNAse H protocol from New England Biolabs (NEB). The sequencing was performed using DNBseq™ technology, targeting approximately 20 million paired-end 100 base pair reads per sample. The reads were aligned to the human reference genome (GRCh38). Quality control of the Fastq files was conducted using MultiQC. Alignment and quantification of the reads were performed using the STAR aligner and the GenCode v33 annotation guide respectively ^40^. This was followed by gene-level read count extraction using FeatureCounts. A filtering process was then applied to remove genes with low expression, those located on sex chromosomes, and non-protein-coding genes to focus the analysis on the most informative transcripts. Normalization of the expression data to control for technical variations was performed using the DESeq2 R package, which adjusts for differences in library size and sequencing depth among samples^41^. Additionally, the removeBatchEffect function from the limma R package was used to correct for potential batch effects associated with RNA integrity number (RIN) values ^42^.

### Mixture Analysis of the Association between Prenatal Pesticide Mixtures and Placental Gene Modules

In this study, we utilized the Weighted Gene Co-expression Network Analysis (WGCNA) approach, as described by Qian et al., to construct and examine networks of co-regulated genes within the placental transcriptome, identifying 21 distinct gene modules ^26^. The biological annotation of these modules, which was performed in their study through functional enrichment analysis using the hypeR R package and the Molecular Signatures Database (MSigDB) to identify hallmark gene sets, provided key insights into the functional roles of the genes ^26^. Additionally, to capture cumulative exposure across pregnancy, we summed the concentrations of the same organophosphate metabolites measured at three distinct time points (early, mid, and late pregnancy). These summed values represented metabolites across pregnancy and were used in our statistical analyses. For the analysis of associations between these placental gene modules and variable exposure levels to prenatal pesticide mixtures, we utilized the quantile g-computation modeling approach available in the quantile g-com R package ^43^. This method facilitated the estimation of scaled effect sizes, which quantify the association of exposures between each module. We adjusted for relevant covariates including maternal age, gestational days, infant sex, early BMI, and location. Certain variables were not examined due to the homogeneity of the participant population, which primarily consisted of individuals from similar backgrounds. Furthermore, to evaluate the variability in gene expression responses to pesticide exposure based on infant sex and specific gestational periods, we conducted stratified analyses, adjusting for the previously mentioned confounders (excluding infant sex in the sex-stratified analysis) ^44^. This involved segmenting the data into subgroups based on the offspring sex and the trimester during which exposure occurred, allowing for a comprehensive examination of timing- and sex-specific effects. Statistical significance for all tests was set at a threshold of p < 0.05.

### Ethical Considerations

All study protocols were reviewed and approved by the Institutional Review Boards of Emory University and the Ethical Review Board of the Institute of Health Sciences at Chiang Mai University. Informed consent was obtained from all participants prior to data collection, in compliance with international ethical standards.

## Results

### Associations Between Prenatal Pesticide Mixture Exposure and Placental Modules

We investigated the relationship between mixed classes of prenatal urinary pesticide metabolites levels and specific placental modules indicative of diverse biological processes. Our mixture analysis revealed significant associations primarily involving two placental modules, both with and without covariate adjustment (Fig 1). The myogenesis module exhibited a notable inverse relationship with average pregnancy pesticide mixtures, as indicated by a significant negative mixture slope parameter (β = −0.012, p = 0.026). The major contributors to the positive effect were DAP (72.3%) and trans-DCCA (27.7%), while DEP (46.5%), DMP (23.0%), and cis-DCCA (17.1%) contributed to the negative direction (Supplemental Fig 1). Conversely, the E2F targets module, which plays a critical role in cell cycle regulation, showed a significant positive association with the average pregnancy urinary metabolite mixture (β = 0.013, p = 0.012) (Fig 1). The largest contributors to the positive effect were urinary levels of DEP (45.8%) and DMP (38.0%) (Supplemental Fig 1). After adjusting for covariates, the overall mixture slope for the myogenesis module remained significant (β = −0.016, p = 0.008), indicating a continued association between the combined metabolite levels and a reduction in the outcome (Fig 1). The primary contributors to the negative effect were urinary levels of DEP (48.9%), cis-DCCA (22.5%), and DMP (21.2%) (Fig 2). On the positive side, urinary DAP levels accounted for 66.5% of the effect, with urinary levels of trans-DCCA contributing 33.5% (Fig 2). Similarly, the mixture effect on the E2F targets module was statistically significant after covariate adjustment (β = 0.013, p = 0.012), suggesting a positive relationship between the exposures and the outcome. As in the unadjusted analysis, urinary levels of DEP (45.8%) and DMP (38.0%) contributed most to the positive direction, while urinary DAP levels (84.1%) had the strongest negative contribution (Fig 2).

**Figure 1.**
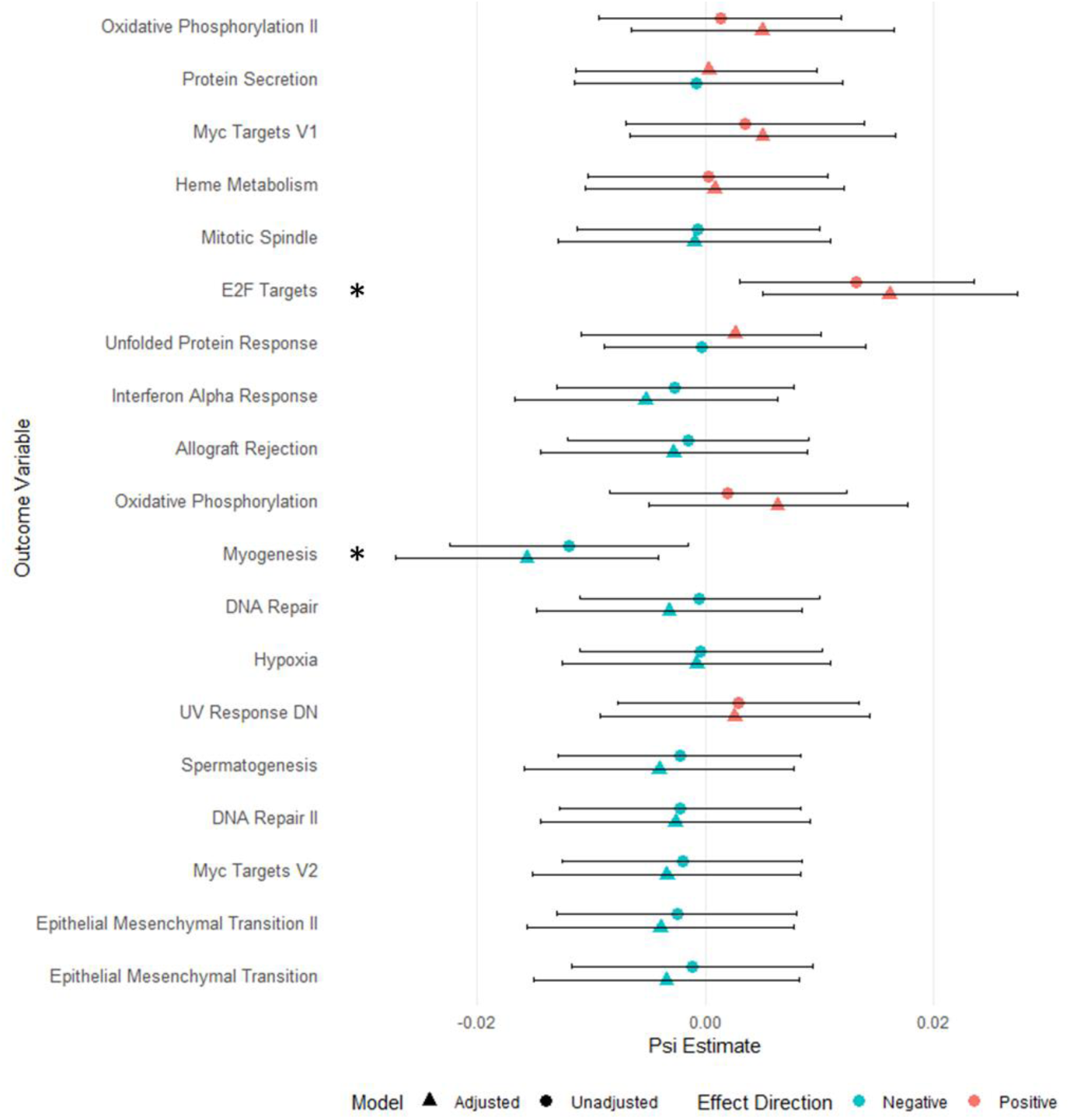
Effect Sizes of Prenatal Urinary Pesticide Metabolite Levels on Placental Gene Modules. This figure presents the effect sizes from quantile g-computation of average pregnancy prenatal pesticide exposure mixtures on various placental gene modules, derived from WGCNA. Effect sizes are shown for both unadjusted (circles) and adjusted (triangles) models. Adjustments were made for covariates including maternal age, gestational days, early pregnancy BMI, infant sex, and study site location (what are the 2 locations?). Each point represents the effect size of a 1 quantile increase in the exposure mixture for a specific module, with red points indicating positive associations and green points indicating negative associations. Error bars represent the confidence intervals for each effect size estimate. Modules marked with an asterisk (*) indicate statistical significance at p < 0.05.

**Figure 2.**
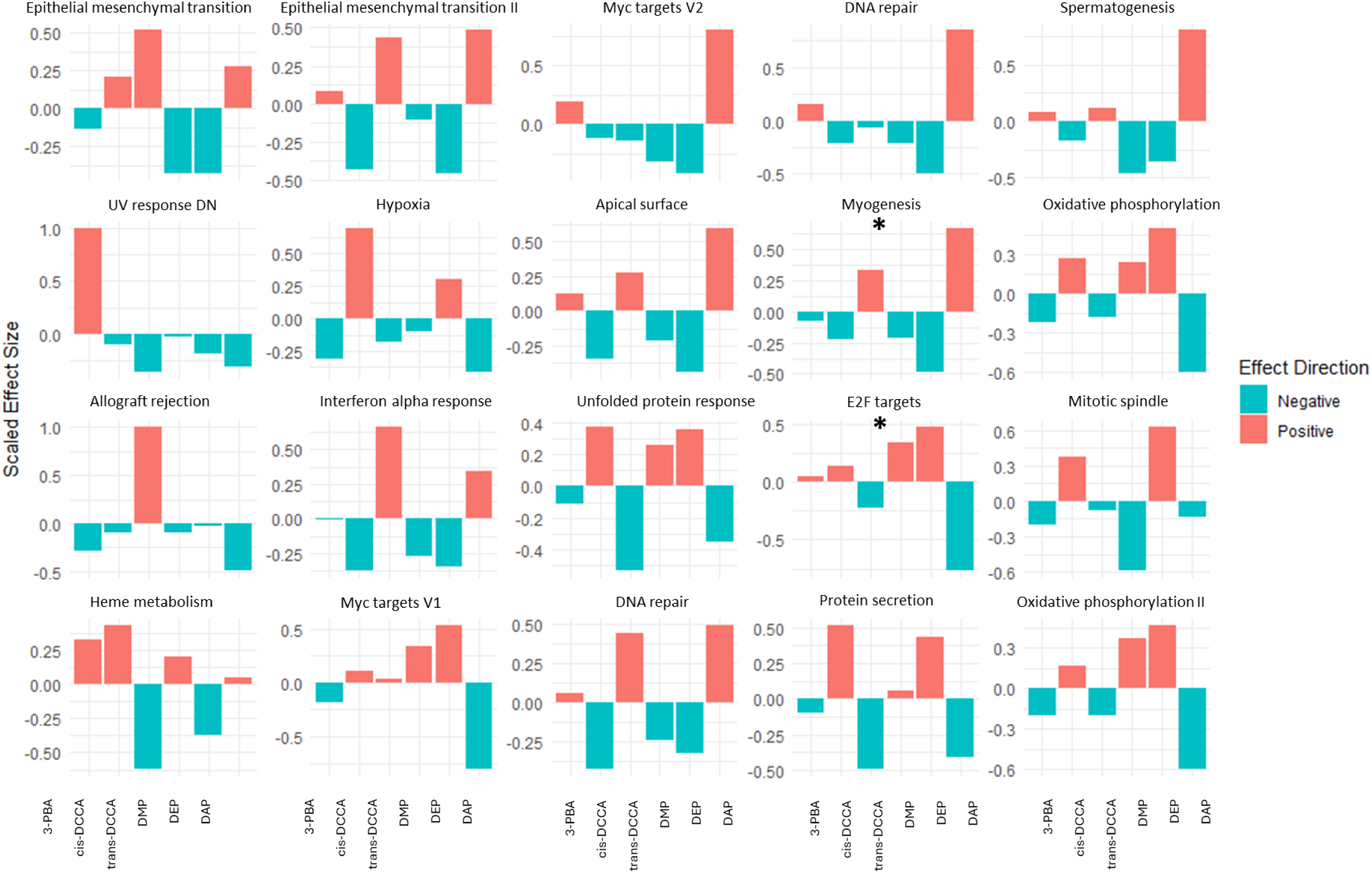
Comprehensive Impact of Urinary Pesticide Metabolite Levels on Placental Gene Modules. This figure provides a detailed comparison of the covariate adjusted, scaled effect sizes of each of the urinary pesticide metabolites in the quantile g-computation modules on each placental gene module. Each panel corresponds to a specific gene module, with bars representing the scaled effect sizes of each pesticide metabolite, including 3-PBA, cis-DCCA, trans-DCCA, DAP, DEP, and DMP. Red bars indicate positive associations between pesticide metabolites and gene module expression, while green bars indicate negative associations. Adjustments were made for maternal age, gestational days, early pregnancy BMI, infant sex, and location. Modules marked with an asterisk (*) indicate statistical significance at p < 0.05.

The other modules displayed either non-significant or marginal associations with varied effect directions (Fig 1). The analysis of scaled effect sizes, allowing for an examination of the specific exposures exhibiting the highest weight in the mixture adjusted model, revealed considerable variability across placental modules (Fig 2). Notably, trans-DCCA and DAP levels were significant contributors to the associations with the myogenesis and E2F targets modules. Although it did not achieve statistical significance, 3-PBA levels were predominantly in association with the UV response module. Larger effect sizes atributable to organophosphate metabolites were observed in the spermatogenesis module, whereas the heme metabolism module was more influenced by pyrethroid metabolites. The apical surface module displayed comparable effect sizes from both pesticide classes. The levels of the six metabolites which represent the excreted end products from exposures to two distinct classes of pesticides, exhibited distinct paterns of driving the associations across the placental modules.

### Sex-Specific Mixture Analysis of Placental Modules

In sex-stratified analysis, we observed that the modules identified as significant prior to stratification— specifically the myogenesis and E2f targets—had notable sex-specific effects (Fig 3). The myogenesis module exhibited a pronounced and significant negative association with average pregnancy exposure mixture in females (β = −0.018, p = 0.022), but this association was not evident in males (β = −0.0028, p = 0.711). Conversely, the E2F targets module showed no significant association in either sex. Although these two modules displayed a consistent patern of association across sex, differences emerged in other areas: 13 out of 20 modules showed opposite effects between sexes, though these were not statistically significant (Supplemental Fig 2). The remaining six modules exhibited similar trends across sex, but with minimal effect sizes that were closely aligned. Furthermore, sex-specific analyses within the same module revealed distinctive paterns in effect sizes (Fig 4). In the myogenesis module, while trends between females and males were similar, the effects were notably more pronounced in females, driven by significant contributions from urinary levels of trans-DCCA and DAP. In contrast, the E2F target module, although not statisticallysignificant, demonstrated markedly different directions of association for urinary metabolites such as cis-DCCA and trans-DCCA between infant sex. For other modules, variations in the direction and magnitude of associations were even more pronounced, highlighting the sex-specific physiological responses to prenatal urinary metabolite levels and underscoring the complexity of these interactions within placental biology.

**Figure 3.**
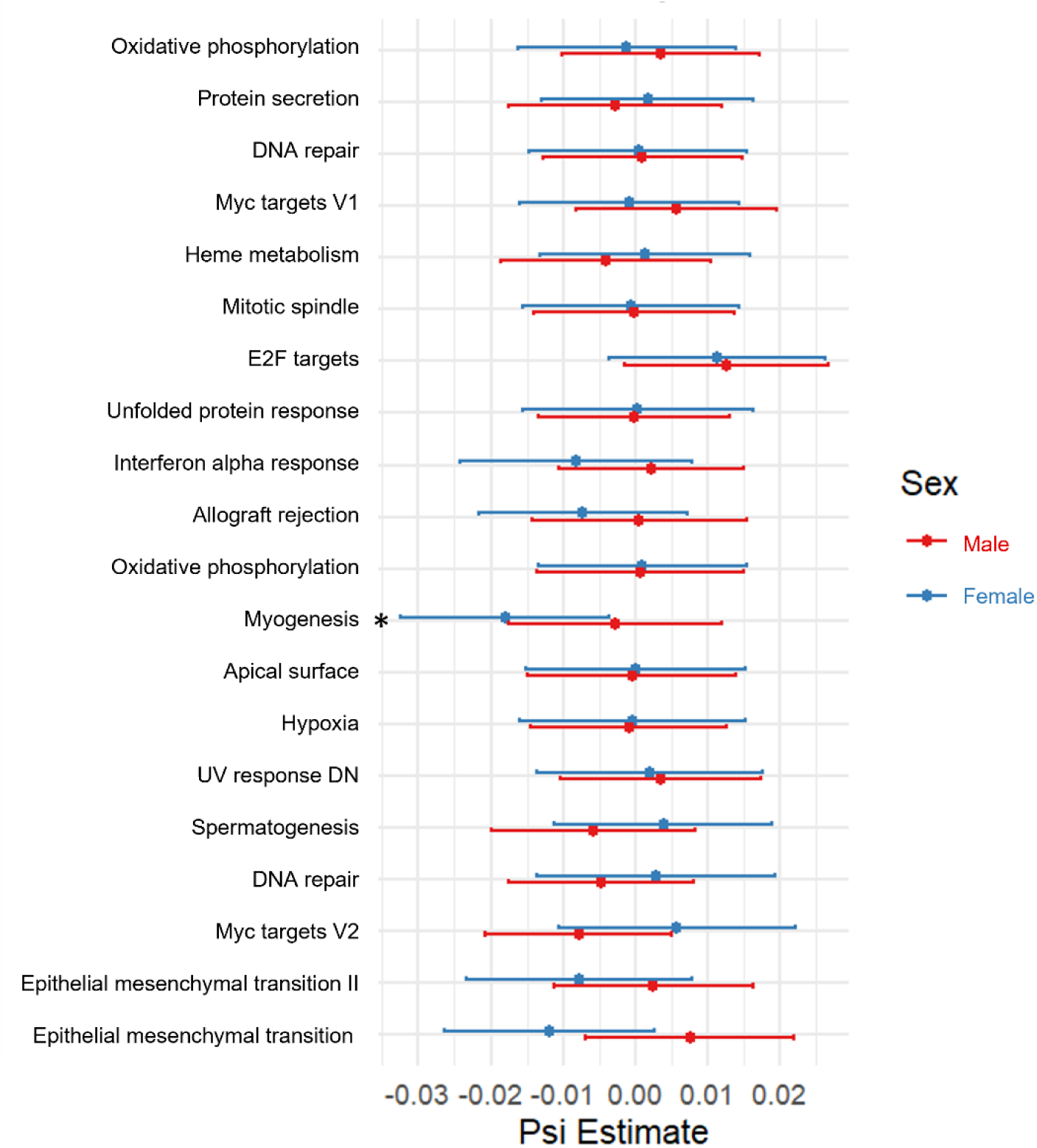
Sex-Specific Effects of Prenatal Urinary Pesticide Metabolite Levels on Placental Gene Modules. This figure illustrates the sex-specific effect sizes from covariate adjusted, quantile g-computation models of average pregnancy prenatal pesticide exposure mixtures on each placental gene module. Red represents male infants, and blue represent female infants. Each line represents the confidence interval for the corresponding effect size estimate. Adjustments were made for maternal age, gestational days, early pregnancy BMI, and study site location (locations). Modules marked with an asterisk (*) are statistically significant at p < 0.05.

**Figure 4.**
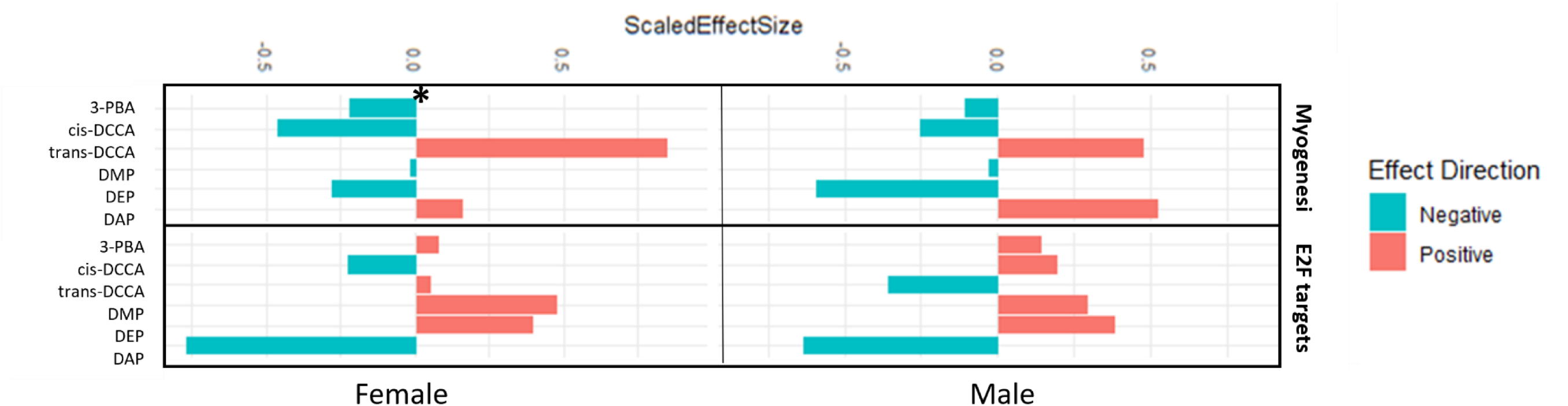
Sex-Specific Responses in Significant Placental Gene Modules to Pesticide Metabolite Mixture Levels. This figure highlights the covariate-adjusted, sex-specific scaled effect zes from covariate adjusted, quantile g-computation models of pesticide metabolites on two key placental gene modules, Myogenesis and E2F Targets. The vertical bars represent the scaled effect sizes for each pesticide metabolite, with red bars indicating positive associations and green bars indicating negative associations. The left panel presents results for female infants, while the right panel displays results for male infants. Adjustments were made for maternal age, gestational days, early pregnancy BMI, and study site location. Modules marked with an asterisk (*) indicate statistical significance p <0.05.

### Trimester-Specific Mixture Analysis of Placental Modules

We also explored the scaled effects of various pesticide metabolite levels on placental modules across three trimesters (Fig 5), focusing particularly on the myogenesis and E2f targets modules (Fig 6). Only the myogenesis were consistently significant during the second and third trimester (Fig 5). The negative overall association between the mixture and the myogenesis module appeared to be driven by associations observed in the middle trimester (β = −0.013, p=0.015) and late trimester (β = −0.012, p=0.028). Urinary levels of trans-DCCA were the predominant driver of the association across all trimesters (76.6%, 81.4%, and 81.9% respectively) (Fig 6). However, urinary levels of DAP and 3-PBA presented minor positive contributions, while levels of metabolites like cis-DCCA, DEP, DMP predominantly displayed negative contributions across all pregnancy periods. For the E2F target module, crucial for cell cycle, the early trimester highlighted a dominant positive contribution from DMP levels (77.6%), contrasting with its drastic shift to a complete negative contribution in the middle trimester. Conversely urinary cis-DCCA and DAP levels shifted from negative to positive contributions between the early and middle trimesters. By the late trimester, the positive contributions were predominated by levels of cis-DCCA, 3-PBA, and DAP, indicating a potential change in the metabolite impact dynamics over time.

**Figure 5.**
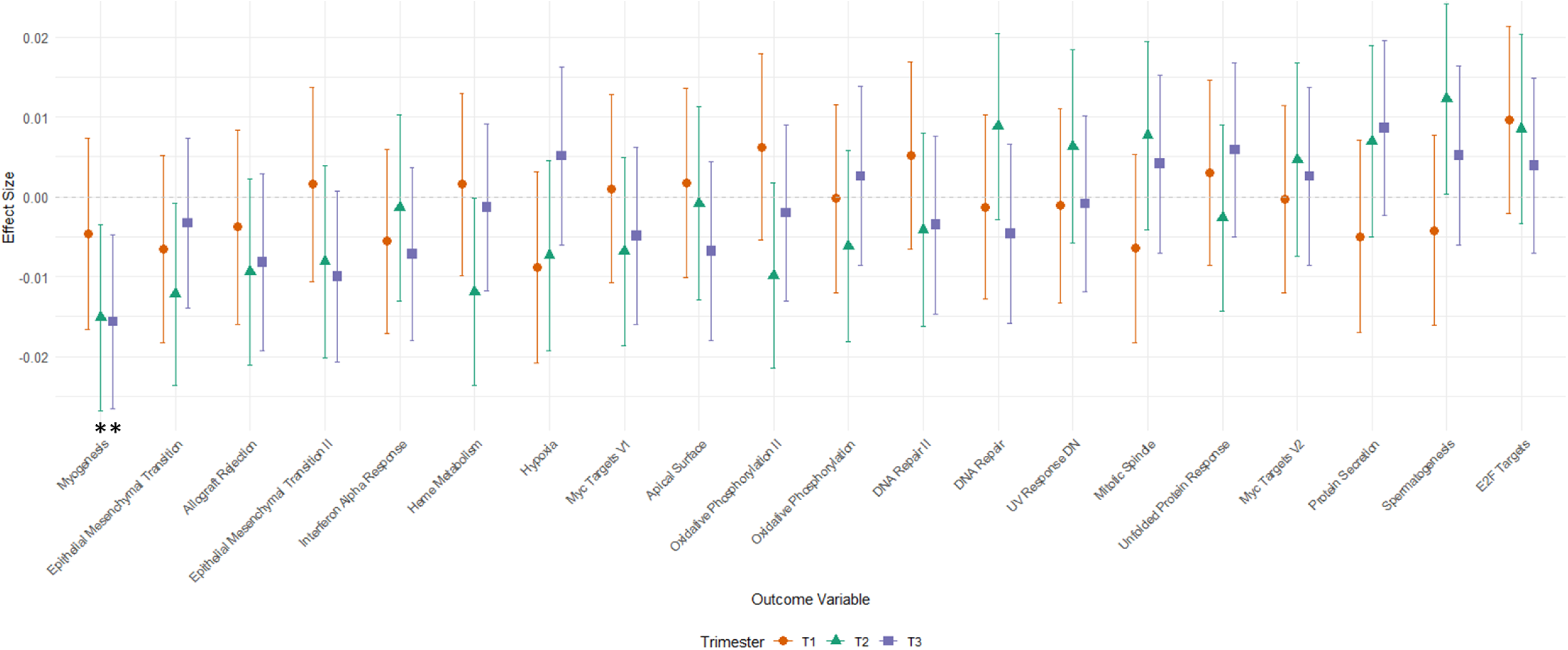
Trimester-Specific Effects of Prenatal Urinary Pesticide Metabolite Levels on Placental Gene Modules. This figure illustrates the trimester-specific effect sizes from covariate-adjusted, quantile g-computation models of prenatal pesticide exposure mixtures on each placental gene module. The colors represent different trimesters, with orange for the first trimester, green for the second trimester, and purple for the third trimester. Each line represents the confidence interval for the corresponding effect size estimate. Adjustments were made for maternal age, gestational days, early pregnancy BMI, infant sex, and study site location. Modules marked with an asterisk (*) are statistically significant at p < 0.05.

**Figure 6.**
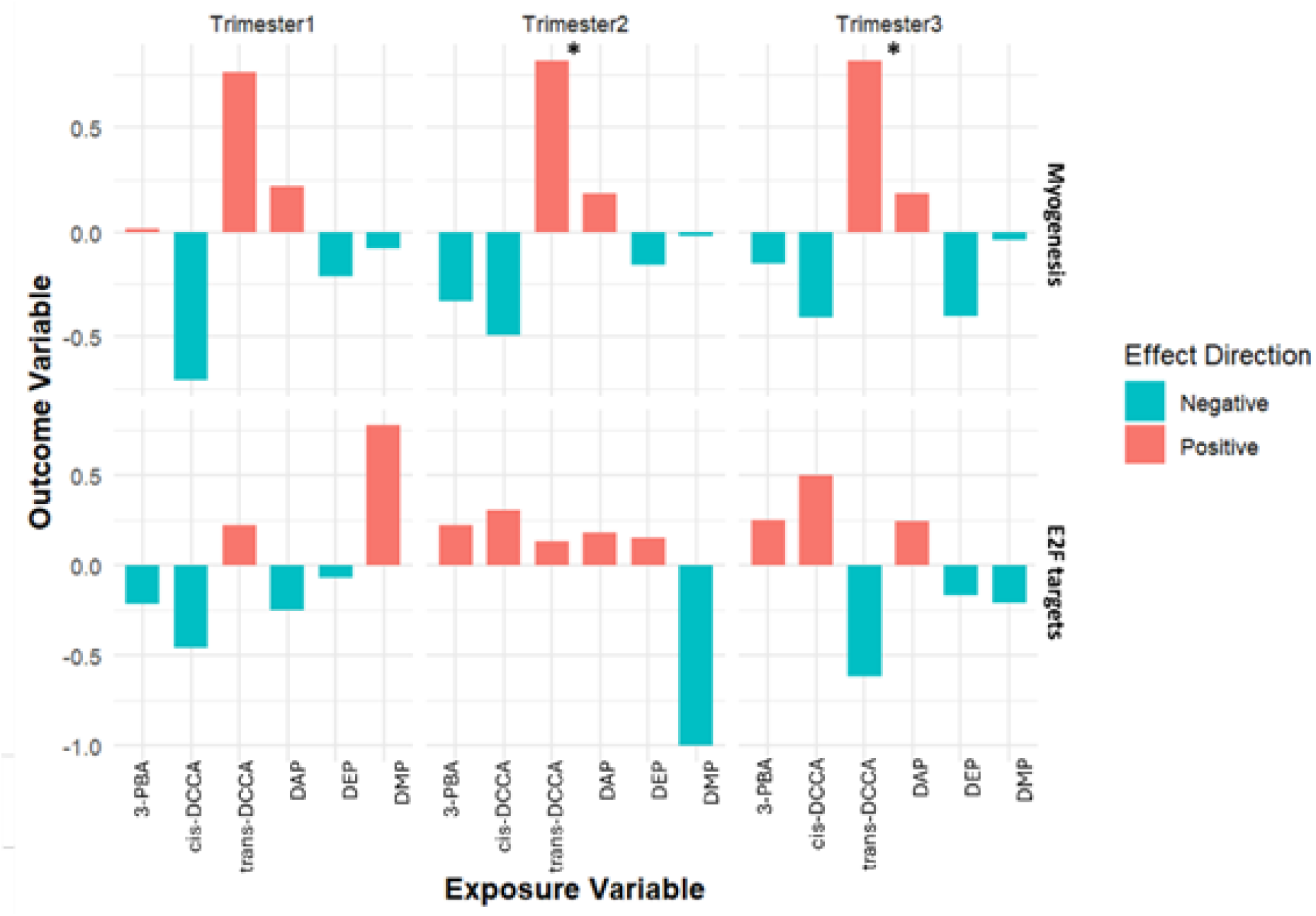
Trimester-Specific Responses in Significant Placental Gene Modules to Urinary Pesticide Metabolite Mixture Levels. This figure highlights the covariate-adjusted, trimester-specific scaled effect sizes from quantile g-computation models of urinary pesticide metabolite exposures (3-PBA, cis-DCCA, trans-DCCA, DAP, DEP, and DMP) on two key placental gene modules: Myogenesis and E2F Targets. The vertical bars represent the scaled effect sizes for each pesticide metabolite across the first, second, and third trimesters of pregnancy, with red bars indicating positive associations and green bars indicating negative associations. Adjustments were made for maternal age, gestational days, early pregnancy BMI, infant sex, and study site location. Modules marked with an asterisk (*) indicate statistical significance at p < 0.05.

## Discussion

Our investigation into the associations between levels of prenatal pesticide metabolite mixtures and placental gene modules revealed a link between the mixtures and the placental myogenesis module, with this association being more pronounced in females and during the middle and late trimesters. Conversely, pesticide metabolite mixture across pregnancy displayed a marginally significant positive association with a module representing E2F targets within the placenta, involved in cell cycle regulation, stronger in males but not pronounced during a specific period of pregnancy. Further analysis of scaled effect sizes across the placental modules underscores the influential roles of levels of metabolites such as trans-DCCA and DMP. While the scaled effects are generally consistent across overall, sex-specific, and trimester-specific analyses for myogenesis, notable dramatic variations in the direction and magnitude of associations between mixtures of six metabolites which represent the excreted end products from exposures to two distinct classes of pesticides are observed in other placental functions.

Our sex-stratified analysis revealed directionally distinct effects across two significant modules, underscoring the complex molecular interactions of pesticides and their varied impacts based on offspring sex. This analysis revealed stronger effect sizes in female infants aligning with existing literature that points to sex-specific vulnerabilities to pesticide exposure. For instance, research by Freire et al. identified male infants as particularly susceptible to endocrine disruption from organochlorine pesticides, while Liu et al. found male infants more affected by neurodevelopmental issues due to organophosphorus pesticides ^45,46^. Conversely, studies also demonstrated significant impacts on birth weight and reproductive hormones, predominantly in male and female infants, respectively ^46,47^. These findings accentuate the necessity of incorporating sex as a factor in environmental risk assessments and illustrate the intricate interplay between biological susceptibilities and environmental exposures. The consistent scaled effects observed across both overall and sex-specific analyses in our study necessitates further investigation into the mechanisms behind these differential responses.

Previous studies from our group focusing on individual classes of pesticides have established foundational insights into their specific impacts on placental function ^21,26,27^. For example, exposure to organophosphates was associated with alterations in placental gene networks crucial for developmental and stress response processes, including pathways involved in epithelial-to-mesenchymal transition and inflammation ^26^. Similarly, late pregnancy levels of DEP were linked to the disruption of gene modules related to myogenesis and EMT in the same study ^26^. The identification of the myogenesis module in the current mixture analysis further supports these findings. This suggests that organophosphate pesticide exposure may interfere with essential processes of muscle development and tissue repair during late gestation, potentially impacting fetal growth and development. In contrast, a study on pyrethroid pesticide metabolites did not reveal direct associations with differential gene expression overall but did identify a negative correlation with genes involved in oxidative phosphorylation, hinting at a subtle impact on energy metabolism in the placenta^27^. These findings delineate the specific effects of single-class pesticides, offering a baseline against which to measure the more complex interactions seen in our current study of mixed pesticide exposures. The present research has uncovered significant modulatory effects within the myogenesis and E2F target modules, underscoring the complex nature of mixed exposures that single-pesticide studies may not fully capture. Notably, these two significant modules exhibited trends over trimesters that suggest dynamic responses to pesticide exposure: a decreasing trend in the myogenesis module and an increasing trend in the E2F target module. Such paterns could indicate an adaptive or resilience response of the placental functions to continuous pesticide exposure. These findings from the current investigation suggest that mixed pesticide exposures may trigger more dynamic and potentially synergistic or antagonistic interactions among the various chemical constituents within the pesticide mixtures. Such complex interactions can profoundly alter biological impacts on the placenta, leading to diverse developmental outcomes for the fetus.

This research examines the placental transcriptome in the context of prenatal pesticide exposures, integrating a diverse array of pesticides to capture the complex interplay of chemical interactions prevalent in agricultural environments. This approach not only broadens our perspective on how such exposures influence placental development and function but also is novel in the application of a mixture analysis framework focusing on the specific transcriptomic responses to prenatal pesticide exposure. Our methodical exploration of the multifaceted nature of environmental risks enhances our capability to develop public health policies and interventions aimed at mitigating adverse prenatal exposures. Additionally, by stratifying data by trimester and infant sex, we provide a detailed examination of how prenatal pesticide exposures differentially affect placental function, enriching our understanding of the temporal and sex-specific susceptibilities to these exposures.

While this study is one of the largest of its kind examining the placental transcriptome in the context of prenatal pesticide exposures, it has several limitations. The sample size, while substantial, does not fully support the extensive range of statistical analyses required, thus constraining our ability to discern subtle variations in gene expression linked to pesticide metabolites. Furthermore, the homogeneity of our cohort may restrict the generalizability of our findings across more diverse populations. To maintain focus during our analyses, we limited the inclusion of confounding variables, which might impact the accuracy of our effect estimates and limit our understanding of causal relationships. Moreover, the use of the quantile g-computation mixture model, while adept at managing the complexities of mixed exposures, complicates our ability to isolate and analyze the effects of individual chemical components within the mixture. This challenge underscores the necessity for future research involving larger, more diverse study populations to comprehensively decipher the complex dynamics of prenatal pesticide exposures and enhance the applicability of our discoveries.

## Conclusion

This study elucidates the complex associations between prenatal urinary pesticide metabolite mixture levels and placental biology, revealing significant variations across sex and trimesters. Notably, the pronounced effects on the myogenesis and E2F target modules suggest their potential as biomarkers for evaluating environmental impacts on placental function and illuminate the potential biological pathways influenced by pesticide mixtures. These findings emphasize the need to reassess how pesticide-related risks are evaluated and highlight the importance of considering the cumulative effects of multiple pesticides in environmental health research and regulatory frameworks. Moving forward, our study sets the stage for future research, highlighting the need for longitudinal studies to assess long-term developmental outcomes in children exposed to mixed pesticides in utero. These studies could help correlate specific placental biomarkers with postnatal health trajectories. Additionally, there is a need to explore intervention strategies to mitigate the risks associated with prenatal pesticide exposure and to understand the specific interactions between different pesticide classes and their cumulative impacts. Such insights are crucial for developing regulatory policies aimed at protecting maternal and fetal health from adverse chemical exposures, contributing to a deeper understanding of the complex dynamics between environmental exposures and placental biology.

## Data availability

Data will be made available on request.

## Acknowledgment

This study was Supported by the US National Institutes of Health (NIH) National Institute of Environmental Health Sciences (NIEHS) grants (R01ES029212, R01ES026082, and P30ES019776). We express our deep gratitude to the participants of the SAWASDEE cohort, as well as the dedicated medical and nursing teams at Fang Hospital.

---

**Table 1.**
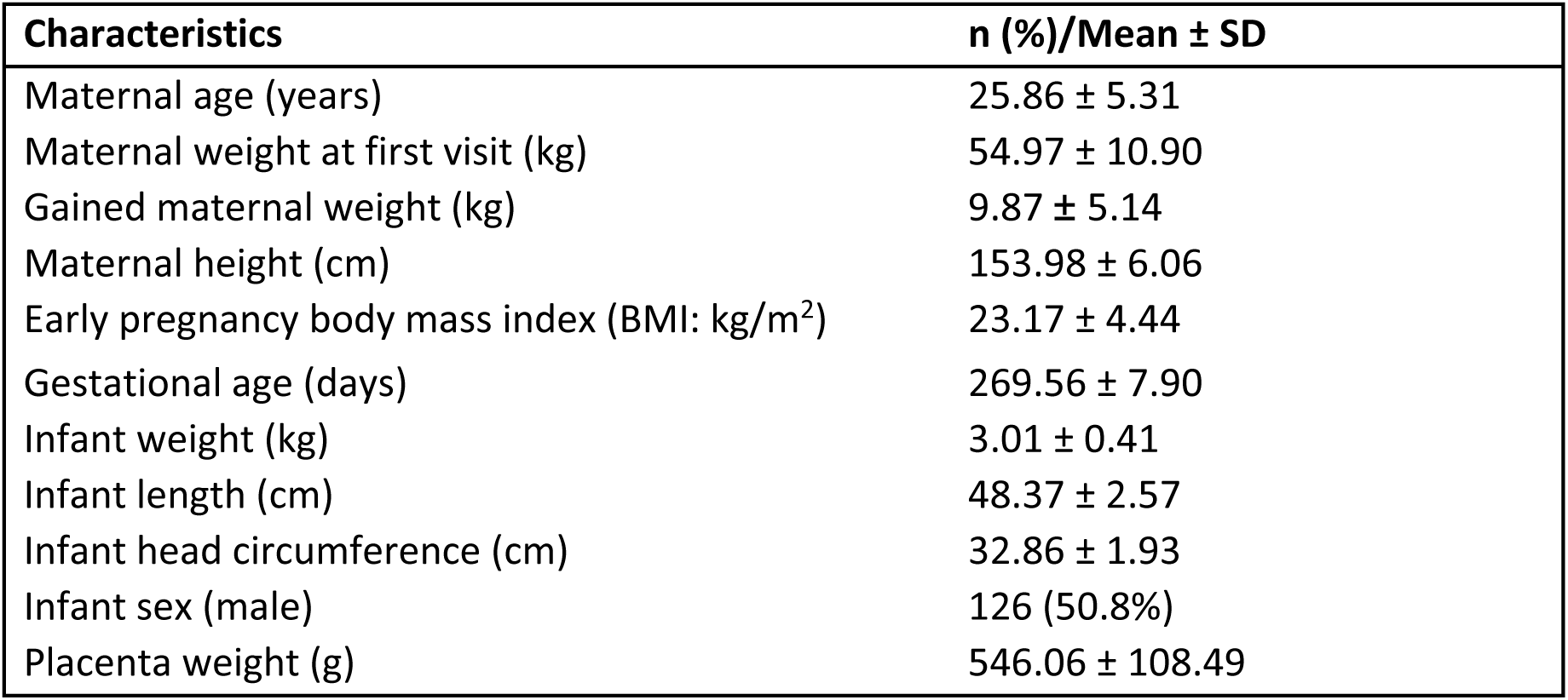
Descriptive characteristics of the study population (N = 248), Study of Asian Women and their Offspring’s Development and Environmental Exposures, 2017–2019.

**Supplemental Figure 1.**
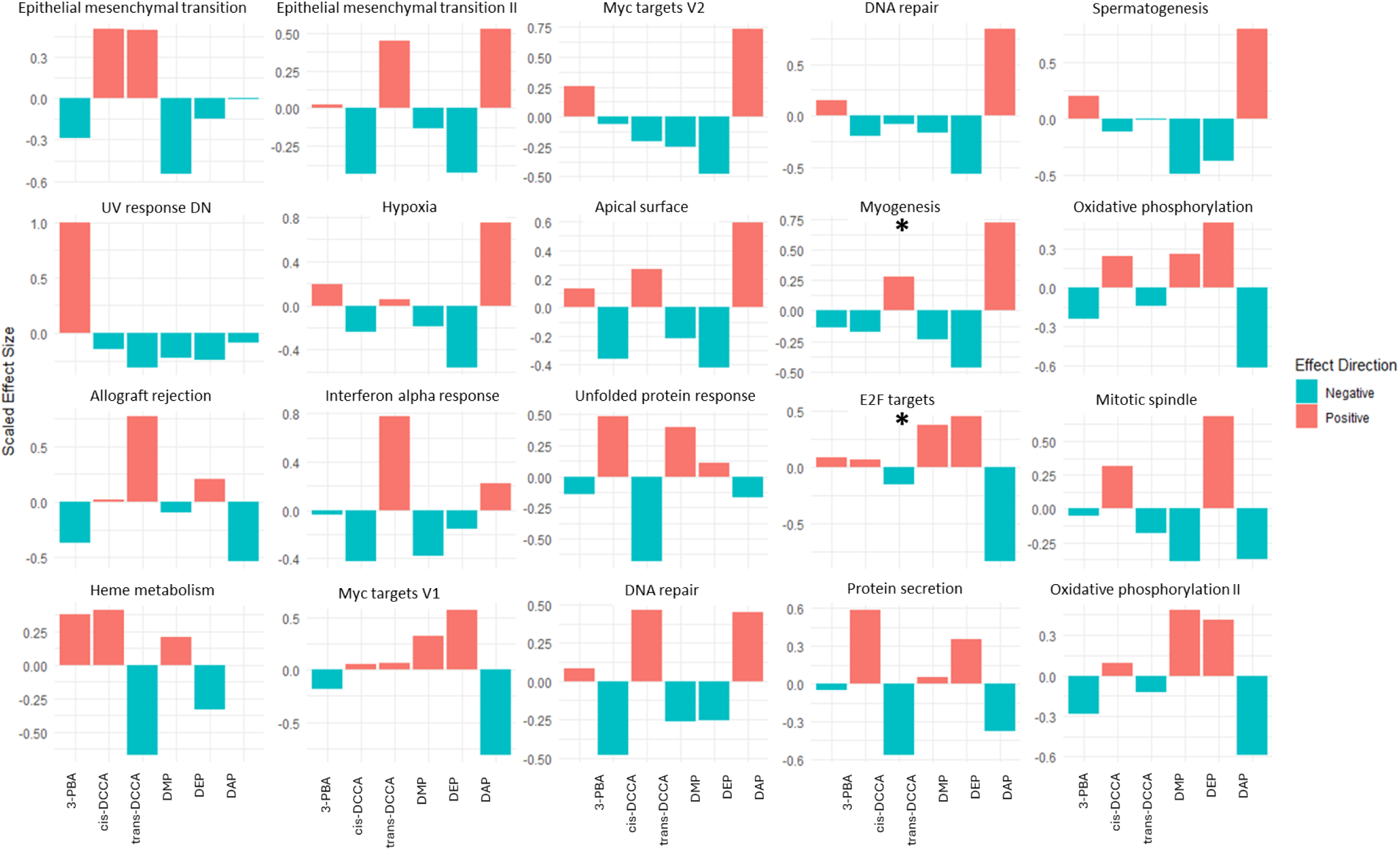
Comprehensive Impact of Urinary Pesticide Metabolite Levels on Placental Gene Modules (Unadjusted Model). This figure presents a detailed comparison of the effects of various pesticide metabolites on multiple placental gene modules in the unadjusted model. Each panel represents a specific gene module, with bars indicating the scaled effect sizes of the pesticide metabolites, including 3-PBA, cis-DCCA, trans-DCCA, DAP, DEP, and DMP. Red bars indicate positive effects, while blue bars indicate negative effects on gene module expression. Error bars represent the confidence intervals for each effect size estimate. Modules marked with an asterisk (*) are statistically significant at p < 0.05.

**Supplemental Figure 2.**
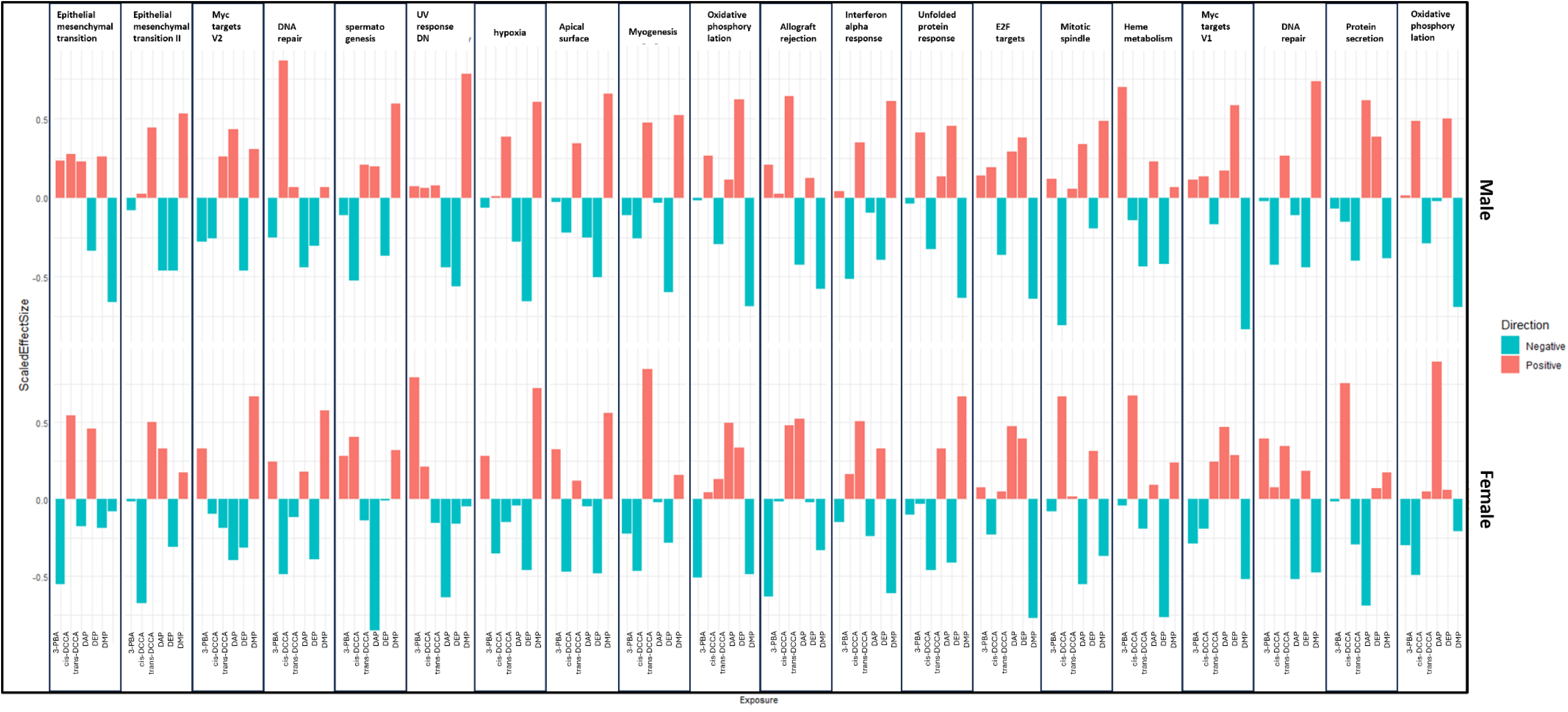
Sex-Specific Impacts of Urinary Pesticide Metabolite Mixture Levels on Placental Gene Modules. This figure illustrates the sex-specific effects of various pesticide exposures on placental gene modules in the adjusted model. Each bar represents the scaled effect size for a specific gene module in response to different pesticide metabolites. Red bars indicate positive effects, while blue bars indicate negative effects. The top panel shows results for male infants, and the bottom panel shows results for female infants. Error bars denote the confidence intervals for each effect size estimate.

